# Determinants of disease prevalence and antibiotic consumption for children under five in Nepal: analysis and modelling of Demographic Health Survey data from 2006 to 2016

**DOI:** 10.1101/2020.07.08.20149153

**Authors:** Charlotte Zheng, Abilasha Karkey, Tianyi Wang, H. Rogier van Doorn, Sonia Lewycka

## Abstract

**Objectives:** Our aims were to examine the geographic, socio-economic and behavioural factors associated with disease and antibiotic consumption in Nepal between 2006 and 2016 and to explore healthcare seeking patterns and the source of antibiotics.

**Methods:** Cross-sectional data from children under five in households in Nepal was extracted from the 2006, 2011 and 2016 Demographic Health Surveys (DHS). Univariate and multivariate analyses were carried out to assess the association of disease prevalence and antibiotic use with age, sex, ecological zone, urban/rural location, wealth index, maternal smoking, use of clean fuel, sanitation, nutrition, access to healthcare and vaccinations.

**Results:** Prevalence of fever, acute respiratory infection (ARI) and diarrhoea decreased between 2006 and 2016, while the proportion using antibiotics increased. Wealth, use of clean fuel, improved toilet sanitation, nutrition and access to healthcare were associated with reduced rates of disease. Those in the highest wealth index use less antibiotics and antibiotic consumption in rural areas surpassed urban regions over time. Health-seeking from the private sector has overtaken government facilities since 2006 with antibiotics mainly originating from pharmacies and private hospitals. Adherence to WHO recommended antibiotics has reduced over time.

**Conclusions:** With rising wealth, there has been a decline in disease prevalence but an increase in antibiotic use with more access to unregulated sources. Understanding antibiotic use and identifying associated behavioural and socio-economic factors may help to inform interventions to reduce inappropriate antibiotic use whilst ensuring access to those who need them.

## Background

Antimicrobial resistance is a global health threat driven by increased and inappropriate use of antibiotics. Antibiotic consumption in low- and middle-income countries (LMICs) is increasing rapidly, converging to rates seen in high-income settings (1). Despite this, relatively little is known about antibiotic use and resistance in these settings due to a lack of surveillance systems, poor regulation and reduced awareness. In particular, there is limited knowledge on community antibiotic use for common childhood infections. Infections such as pneumonia and diarrhoea remain major global burdens of disease in under-fives (U5) (2) and represent a significant driver of antibiotic consumption.

Nepal is a landlocked country in South Asia with a population of around 30 million. It is one of the poorest countries in the world with 21.6% of the population living below the poverty line in 2018 (3). However the economy is developing rapidly; Kathmandu is the fastest growing metropolitan area in South Asia (4) and the proportion of people living below the poverty line has declined substantially since 2003 (5). The recent increase in population wealth may be partially explained by the authorisation of labour emigration by the Home and Labour ministry in 2011. Those returning from working abroad have been able to afford to move from rural villages to urban areas with greater access to basic necessities. Nepal’s ecology is composed of flat lands (terai), hills and mountain areas making delivery and access to healthcare challenging as well as contributing to disease incidence. Natural disasters affect public health and the ability to provide robust healthcare. The terai experienced massive floods in 2012, 2014 and 2017 leading to the spread of communicable diseases as sanitation and clean water supplies were disrupted. The April 2015 earthquake caused widespread devastation in Nepal and major disruption of healthcare infrastructures.

Total under-five mortality in Nepal was estimated at 32 per 1000 live births (globally it was 39 per 1000 live births) in 2018 with pneumonia, neonatal asphyxia and diarrhoeal diseases among the leading causes (6, 7). The majority of healthcare is community based with primary health care centres and community health volunteers serving as the first port of call particularly in hard to reach locations (8). Recent years have seen a substantial increase in the number of private hospitals and clinics in urban areas, likely reflecting the demands of an increasingly wealthy population.

The Global Antibiotic Resistance Partnership situation analysis for Nepal reports that antibiotics are the most frequently prescribed medication, used both prophylactically and therapeutically and can be purchased routinely in the community from pharmacies, drug stores and informal drug sellers (7). There are no antibiotic stewardship guidelines and little education of antibiotic resistance amongst health professionals (7). Literature on the demographic and behavioural determinants of disease and antibiotic use for common childhood illnesses in Nepal is limited.

The Demographic Health Survey (DHS) provides data on the prevalence and management of childhood acute respiratory infection (ARI), diarrhoea and fever and the social, demographic and economic characteristics of the households surveyed. The data has not previously been used to examine determinants of childhood illness and associated antibiotic consumption. This information could inform health policy and methods in rationalising antibiotic use to reduce resistance while ensuring access to those in most need

## Aims and objectives

Our aims were to examine the geographic, socio-economic and behavioural determinants of ARI, fever and diarrhoea; examine antibiotic consumption at the disease and population level; and identify demographic and behavioural determinants of antibiotic consumption. We also wanted to explore healthcare seeking patterns and the source of antibiotics. By analysing data captured between 2006 and 2016, we sought to explore how behaviours have changed over time.

## Methods

### Datasets

Cross-sectional data on living children under five in households in Nepal were extracted from the 2006, 2011 and 2016 DHS surveys through datasets and survey reports. The DHS survey collects data using a stratified 2 stage cluster sampling method. The time frame for data collection was from February to August in 2006, February to June in 2011 and June to January in 2016. This mostly covered the wet seasons (June – August) when febrile and diarrhoeal illnesses usually peak in incidence.

### Definitions

ARI was defined as fast and/or difficulty breathing due to a problem in the chest with or without cough in the 2 weeks preceding the survey. This is concordant with the definition given by the WHO Integrated Management of Childhood Illnesses (IMCI) for pneumonia. Fever (parameters not specified in the survey) and diarrhoea (frequent loose or liquid stools) were defined as occurrence of the symptoms in the last 2 weeks. Dysentery was ascertained as the presence of diarrhoea with bloody stools. Occurrence of these symptoms was based on maternal/care-giver recall. Care seeking was defined by whether the mother sought advice or treatment for the illness from any source. Antibiotic treatment was assessed by asking the mother if the child had taken any drugs during the illness and if so whether this consisted of antibiotic pills, syrups or injections. Rates of antibiotic use were calculated with the total under-five population as the denominator to reflect antibiotic consumption at the population level (see *Supplementary table 1* in the appendix for a full list of definitions)

**Table 1.** Background characteristics of U5s alive at the time of the survey. Numbers represent weighted numbers in each category to enable comparison between years.

|  | 2006 | 2011 | 2016 |
| --- | --- | --- | --- |
| **U5 population** | 5252 | 5140 | 4887 |
| **Male** | 2681 (51.0%) | 2649 (51.5%) | 2563 (52.4%) |
| **Female** | 2571 (49.0%) | 2490 (48.4%) | 2324 (47.6%) |
| **Age (months)** |  |  |  |
| <6 | 484 (9.2%) | 531 (10.3%) | 423 (8.7%) |
| 6-11 | 494 (9.4%) | 491 (9.1%) | 476 (9.7%) |
| 12-23 | 984 (18.7%) | 1000 (19.4%) | 1029 (21.1%) |
| 24-35 | 1147 (21.8%) | 1013 (19.7%) | 928 (19.0%) |
| 36-47 | 1049 (20.0%) | 1106 (21.5%) | 970 (19.8%) |
| 48-59 | 1094 (20.8%) | 999 (19.4%) | 1060 (21.7%) |
| **Urban** | 652 (12.4%) | 483 (9.4%) | 2649 (54.2%) |
| **Rural** | 4600 (87.6%) | 4656 (90.1%) | 2238 (45.8%) |
| **Mountain** | 443 (8.4%) | 400 (7.8%) | 342 (7.0%) |
| **Hill** | 2171 (41.3%) | 2033 (39.6%) | 1857 (38.0%) |
| **Teraï** | 2638 (50.2%) | 2707 (52.7%) | 2688 (55.0%) |
| **Wealth index** |  |  |  |
| Poorest | 1328 (25.3%) | 1322 (25.7%) | 1041 (21.3%) |
| Second | 1117 (21.3%) | 1212 (23.6%) | 1028 (21.0%) |
| Middle | 1053 (20.0%) | 1071 (20.8%) | 1087 (22.2%) |
| Fourth | 950 (18.1%) | 899 (17.5%) | 999 (20.4%) |
| Richest | 804 (15.3%) | 726 (14.1%) | 732 (15.0%) |
| **Mother's education** |  |  |  |
| No education | 3129 (59.6%) | 2410 (46.9%) | 1663 (34.0%) |
| Primary | 957 (18.2%) | 1032 (20.1%) | 981 (20.1%) |
| Secondary | 1038 (19.8%) | 1411 (27.5%) | 1564 (32.0%) |
| Higher | 128 (2.4%) | 287 (5.6%) | 679 (13.9%) |

### Analysis

A descriptive analysis was carried out on the survey reports and datasets to identify changes from 2006 to 2016 with regards to demographics, disease prevalence and antibiotic use. The source of antibiotics and appropriate use as per the IMCI guidelines were also examined. A univariate analysis was performed to evaluate the association of the following variables with disease prevalence and antibiotic use: age, sex, ecological zone, urban/rural location, wealth index, maternal smoking, use of clean fuel, sanitation, nutrition, access to healthcare and vaccinations. The multivariate model adjusted for age, wealth and urban/rural location. Sample weights provided by the DHS data were applied to account for over and under sampling of particular regions, and adjustments were made for clustering of data using Taylor-linearized variance estimation. Data management and analysis was carried out using Stata version SE 12. All data was publicly available and anonymised with ethical approval covered under the original data collection.

## Results

### Background characteristics

5457, 5054 and 4861 children (unweighted data) were included in the 2006, 2011 and 2016 surveys respectively. The background characteristics of the population studied are summarised in *Table 1*. There were 14 sampling strata for 2016 and 13 for 2006 and 2011 (9-11). Male and female children were equally represented and numbers within each age group were evenly spread across the surveys. The majority of the population sampled lived in the terai region, representative of the general population. The proportion of mothers achieving secondary and higher level education increased from 2006 to 2016. There was a higher representation of urban populations in 2016 which reflects increasing urbanisation and updated urban-rural classifications by the National Population and Housing Census (11).

### Prevalence of childhood illness

Overall, the reported percentage prevalence of disease decreased from 5.9% to 2.4% and from 11.9% to 7.6% between 2006 and 2016 for ARI and diarrhoea, respectively. Conversely, the reported prevalence of fever increased over this time (*Figure 1A*). The peak prevalence of illness was in those aged between 6-23 months.

**Figure 1.**
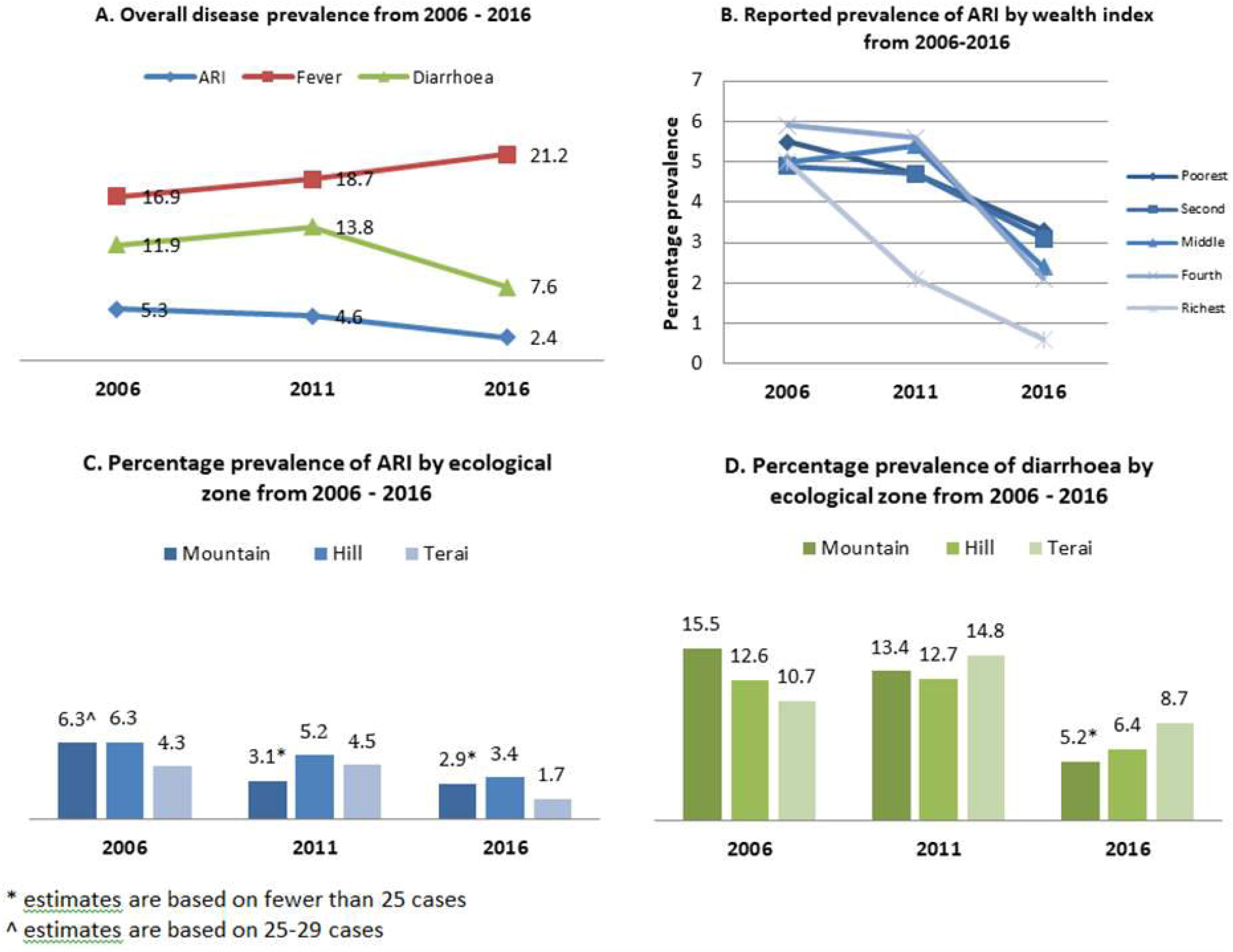
Demographic determinants of disease prevalence from 2006-2016

The prevalence of ARI was highest in the hill areas in all 3 surveys (*Figure 1C*). The largest reductions in ARI prevalence were seen in the highest wealth quintiles, particularly for 2011 and 2016 (*Figure 1B*). Diarrhoea prevalence was lowest in the terai region for 2006, but highest in 2011 and 2016, corresponding to the massive floods in the terai (*Figure 1D*). On the other hand, there was no difference observed between disease prevalence and urban and rural locations. Higher levels of maternal education were associated with increased reported prevalence of fever in 2006 and 2011, although this effect was not seen in the adjusted analysis for 2011. No association was found for maternal education and prevalence of ARI or diarrhoea (*Supplementary Table 3, 5 and Table 2*).

**Table 2.**
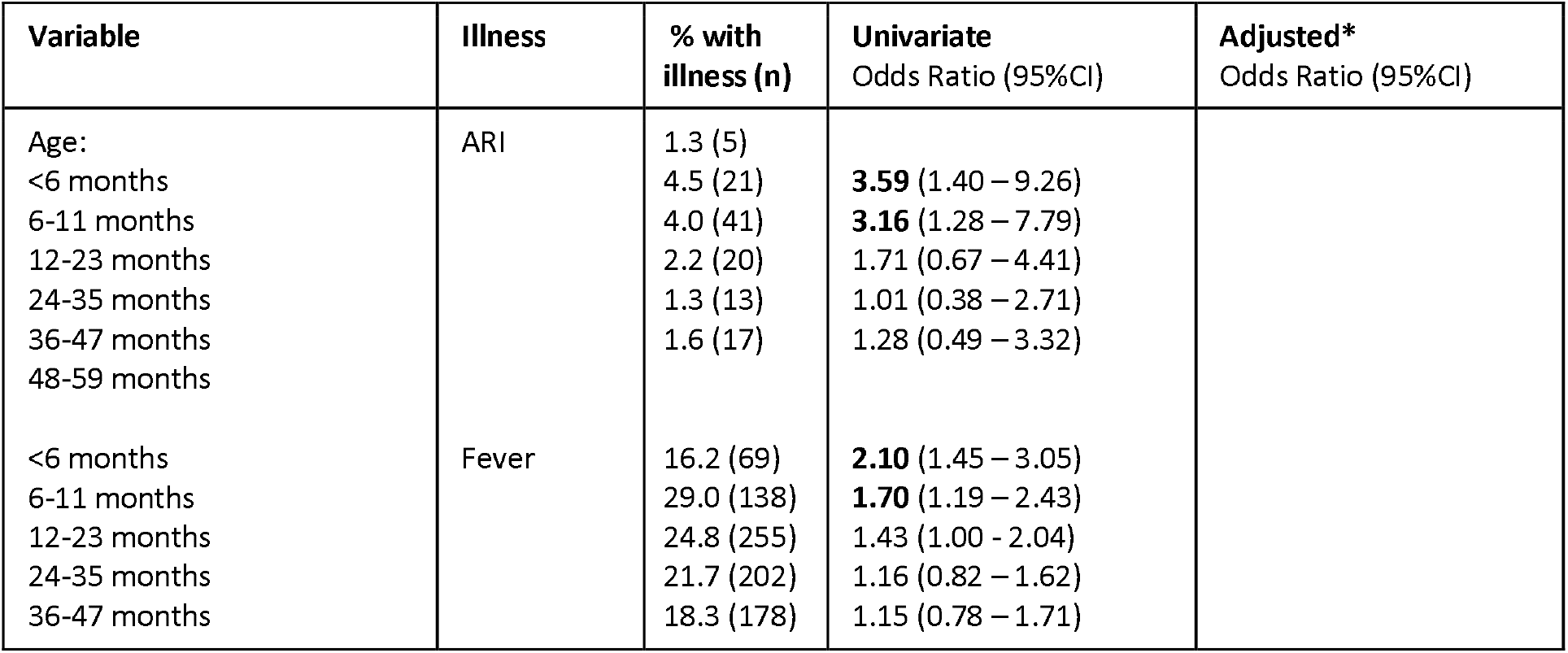
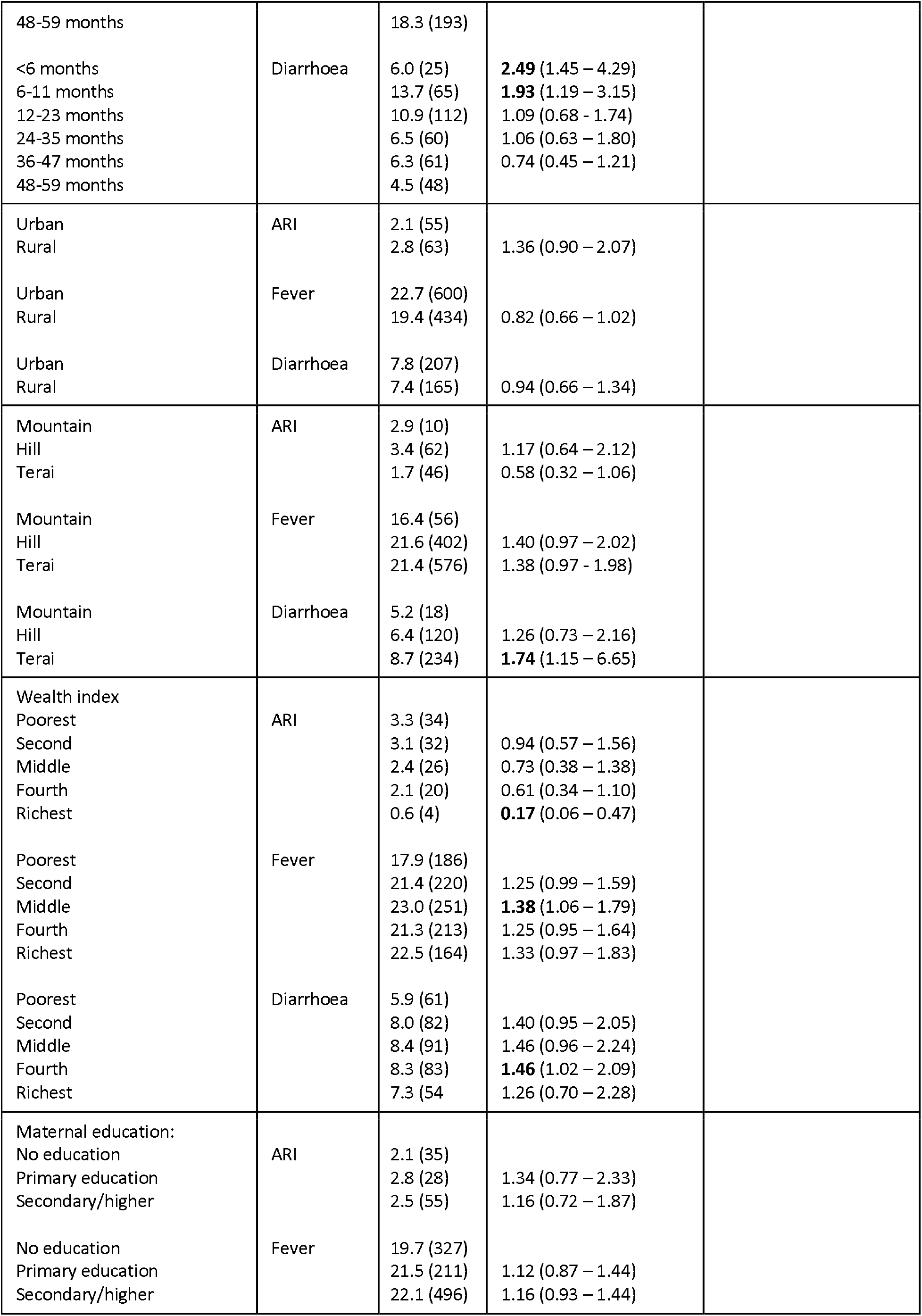
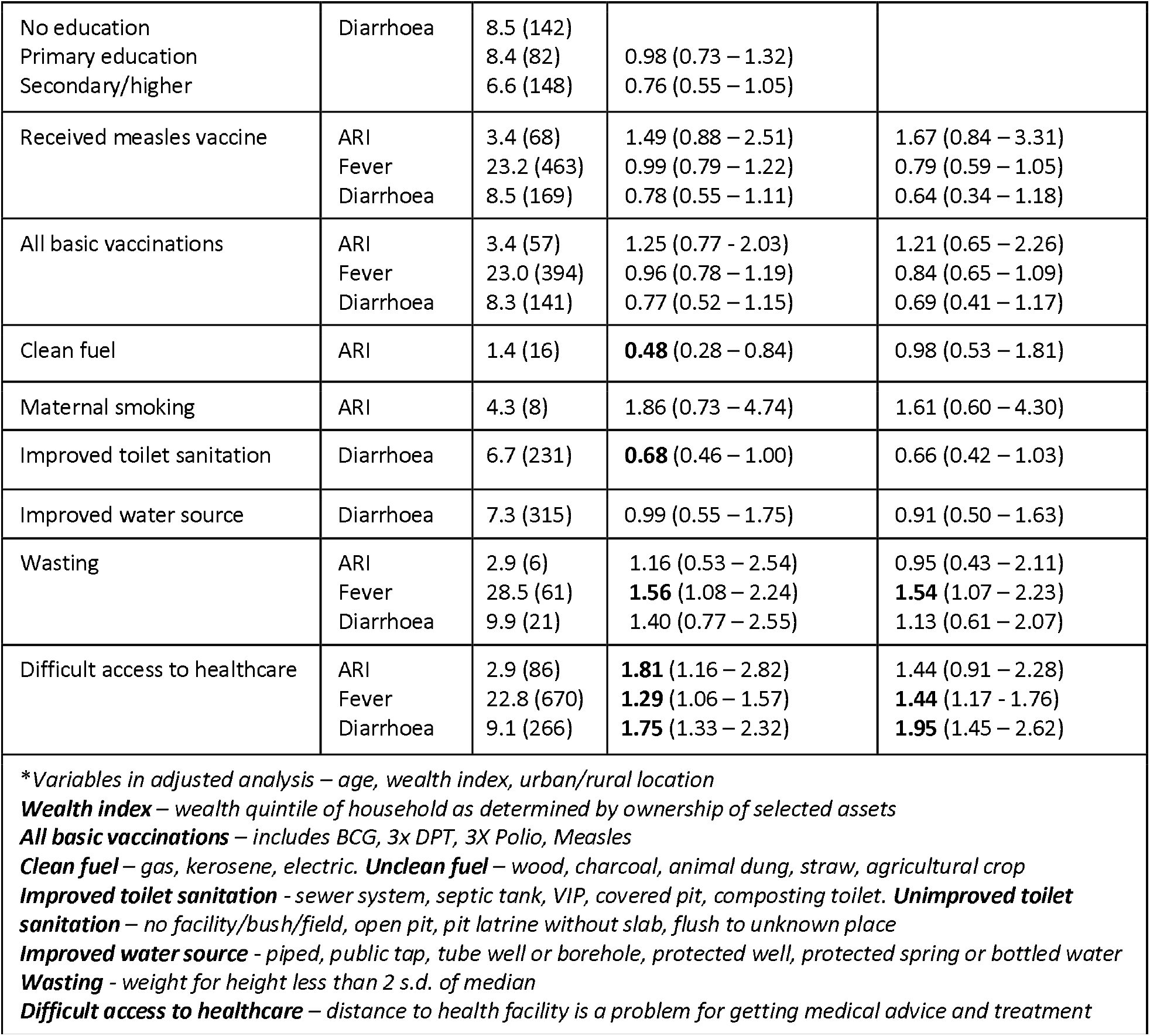
Demographic and behavioural determinants of disease (2016 data)

**Table 3.**
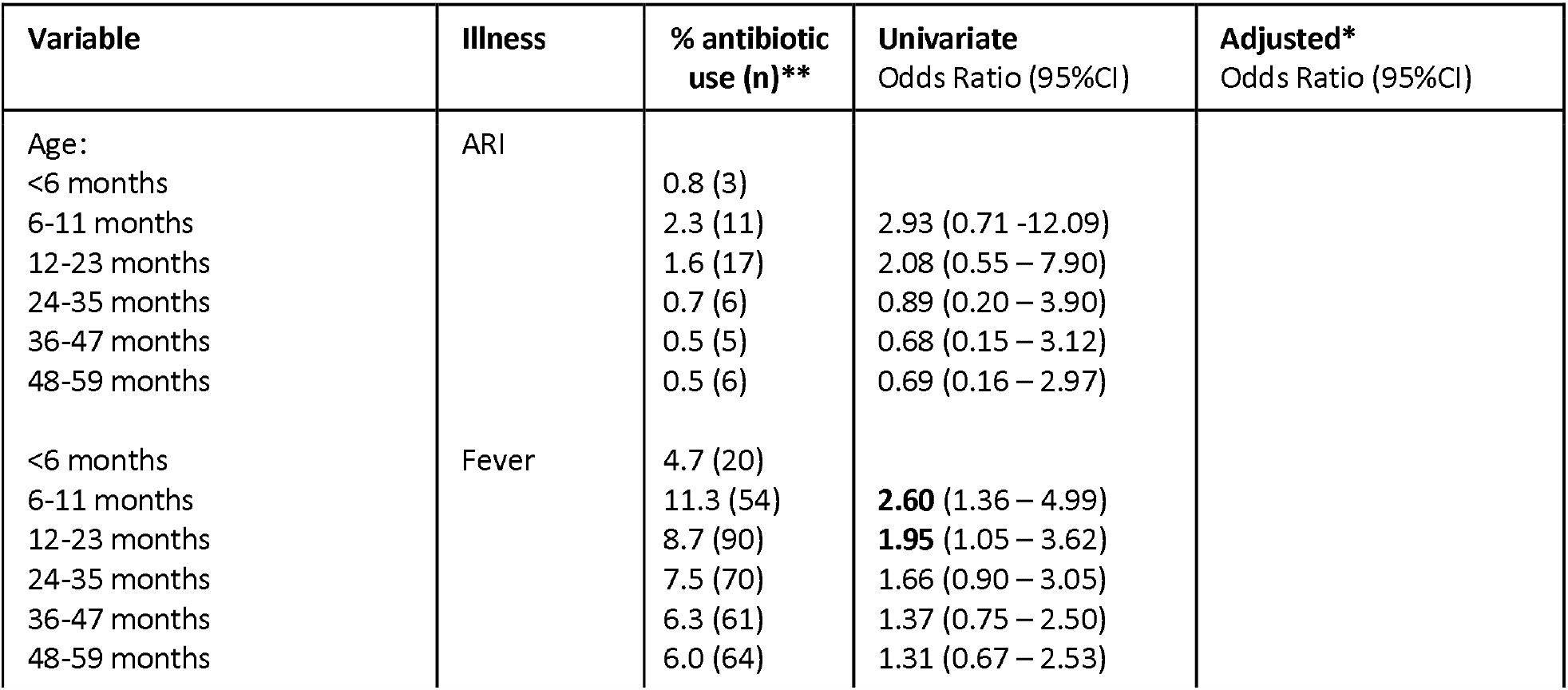
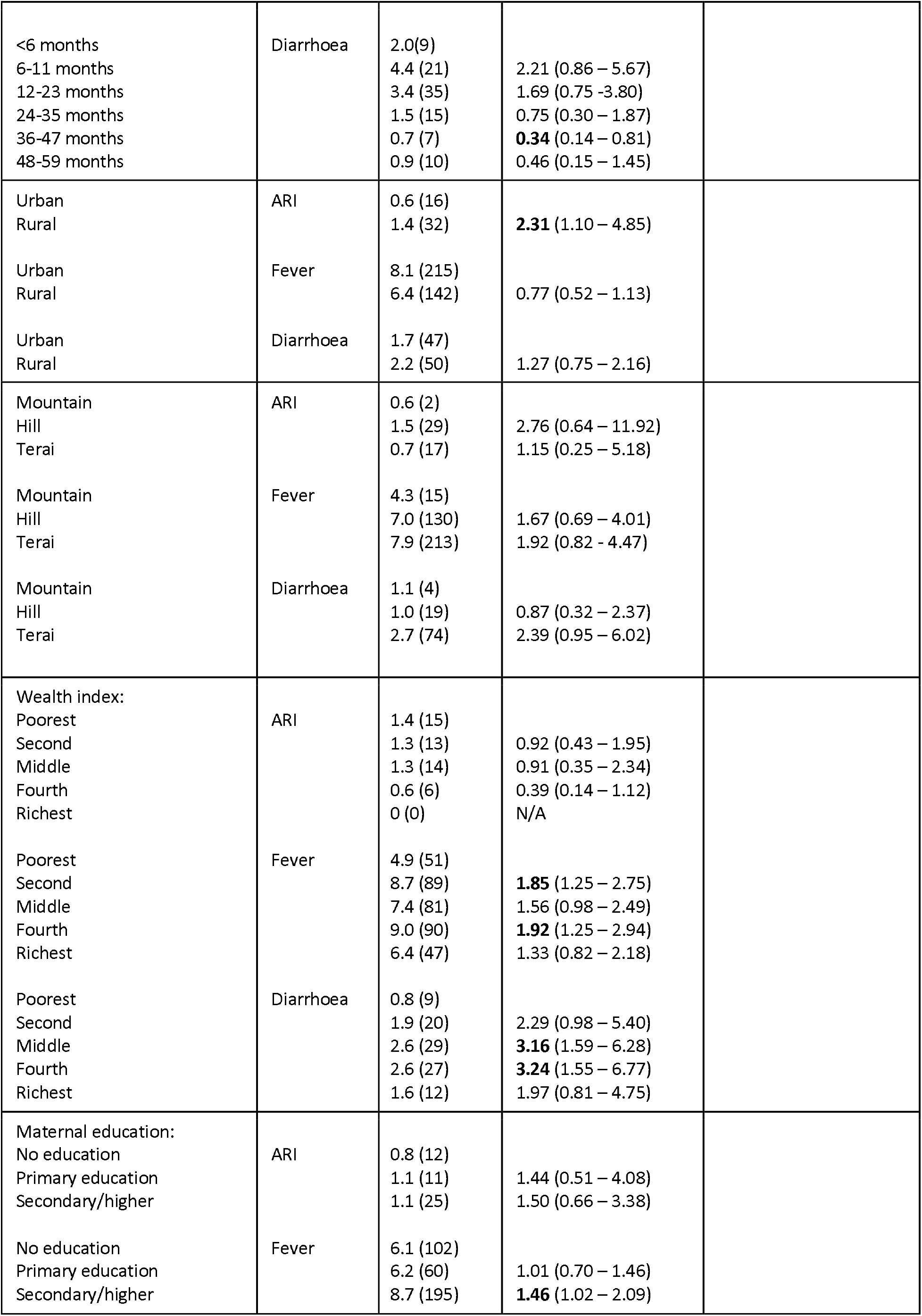
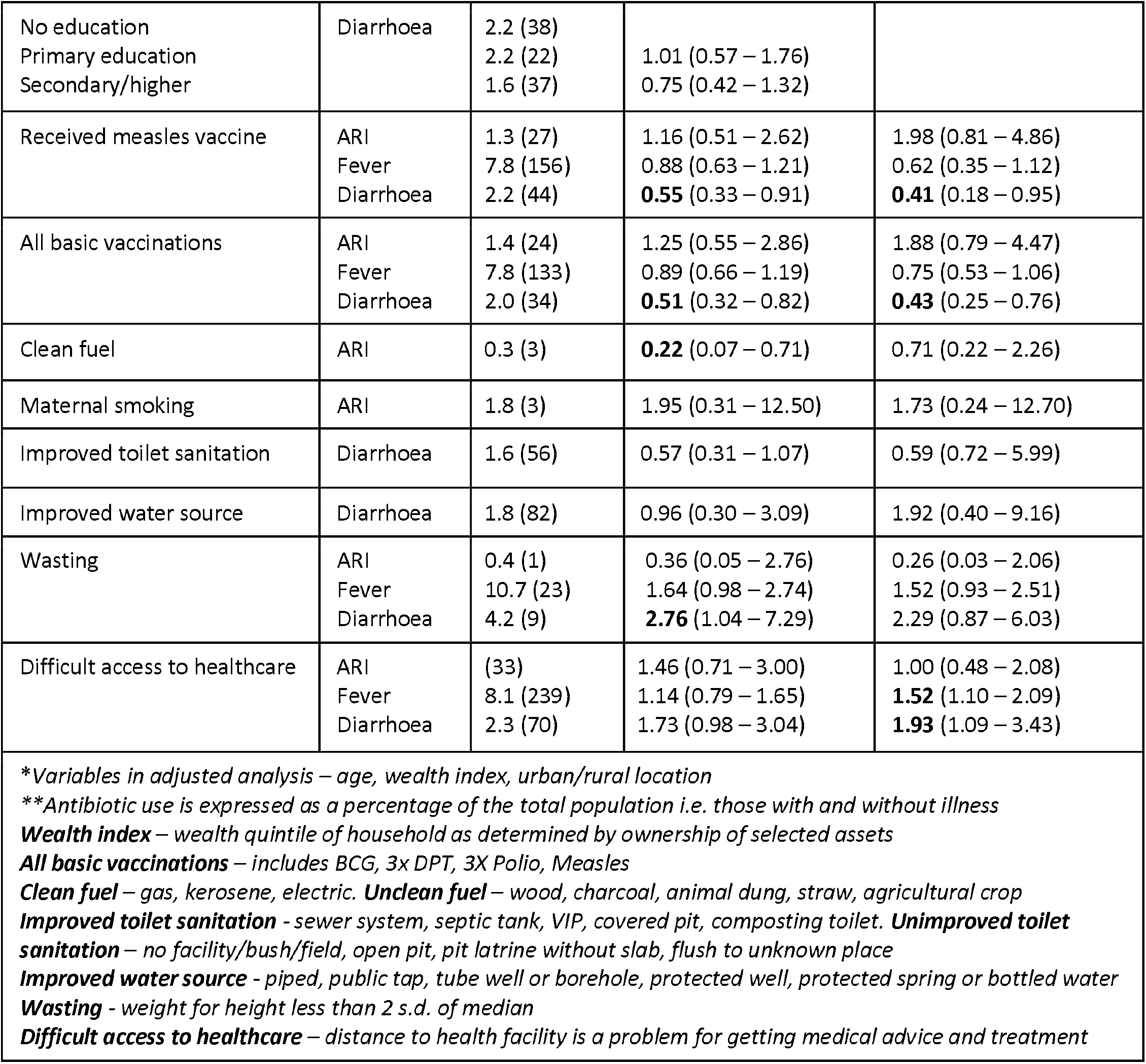
Demographic and behavioural determinants of antibiotic use (2016 data)

In terms of behavioural determinants of disease, clean fuel and improved toilet sanitation were associated with reduced prevalence of ARI and diarrhoea in 2011 and 2016, respectively (*Supplementary Table 5 and Table 3*). This effect was attenuated in the multivariate analysis for clean fuel. Measles vaccination was associated with reduced rates of diarrhoea in 2006 and 2011, although this was not observed in the adjusted model (*Supplementary Table 3 and 5*). Wasting (a proxy for nutritional status) was associated with increased rates of fever and diarrhoea. Difficulty accessing healthcare was associated with higher ARI, fever and diarrhoea prevalence in 2016. There was no association found between disease prevalence and maternal smoking, improved water source, and other vaccinations (*Table 2*).

### Antibiotic use

Despite an overall reduction in ARI and diarrhoea prevalence, the reported antibiotic use increased for all 3 conditions from 2006 to 2016 (*Figure 2A*). In contrast to urban areas, antibiotic use in rural areas increased during each time interval with 2016 data demonstrating rural antibiotic consumption for ARI and fever surpassing those in urban regions (*Figure 2C*). Whilst there is a trend for increasing antibiotic use with increasing wealth for 2006, the 2016 data suggest a reduction in antibiotic use for the wealthiest quintile, which is particularly marked in ARI (*Figure 2B*). Antibiotic consumption for diarrhoea was highest in the middle and fourth wealthiest quintiles for diarrhoea in 2016, whilst rates were highest in the wealthiest quintile in 2006 and 2011. Higher levels of maternal education were generally associated with increased antibiotic use, particularly for fever, where the effect was present in all 3 time periods (*Table 3 and Supplementary Table 4*).

**Figure 2.**
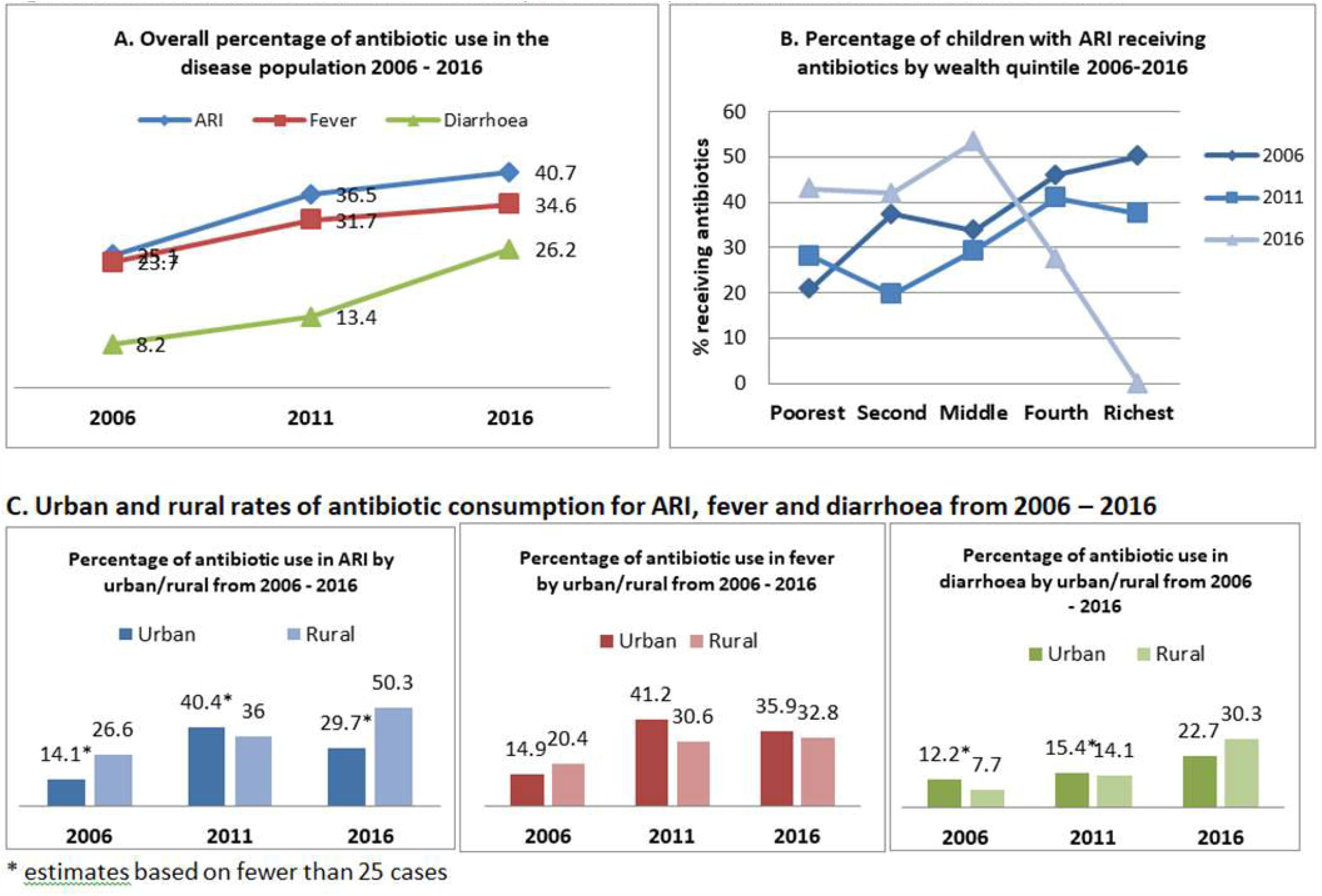
Determinants of antibiotic consumption for ARI, fever and diarrhoea from 2006-2016

**Figure 3.**
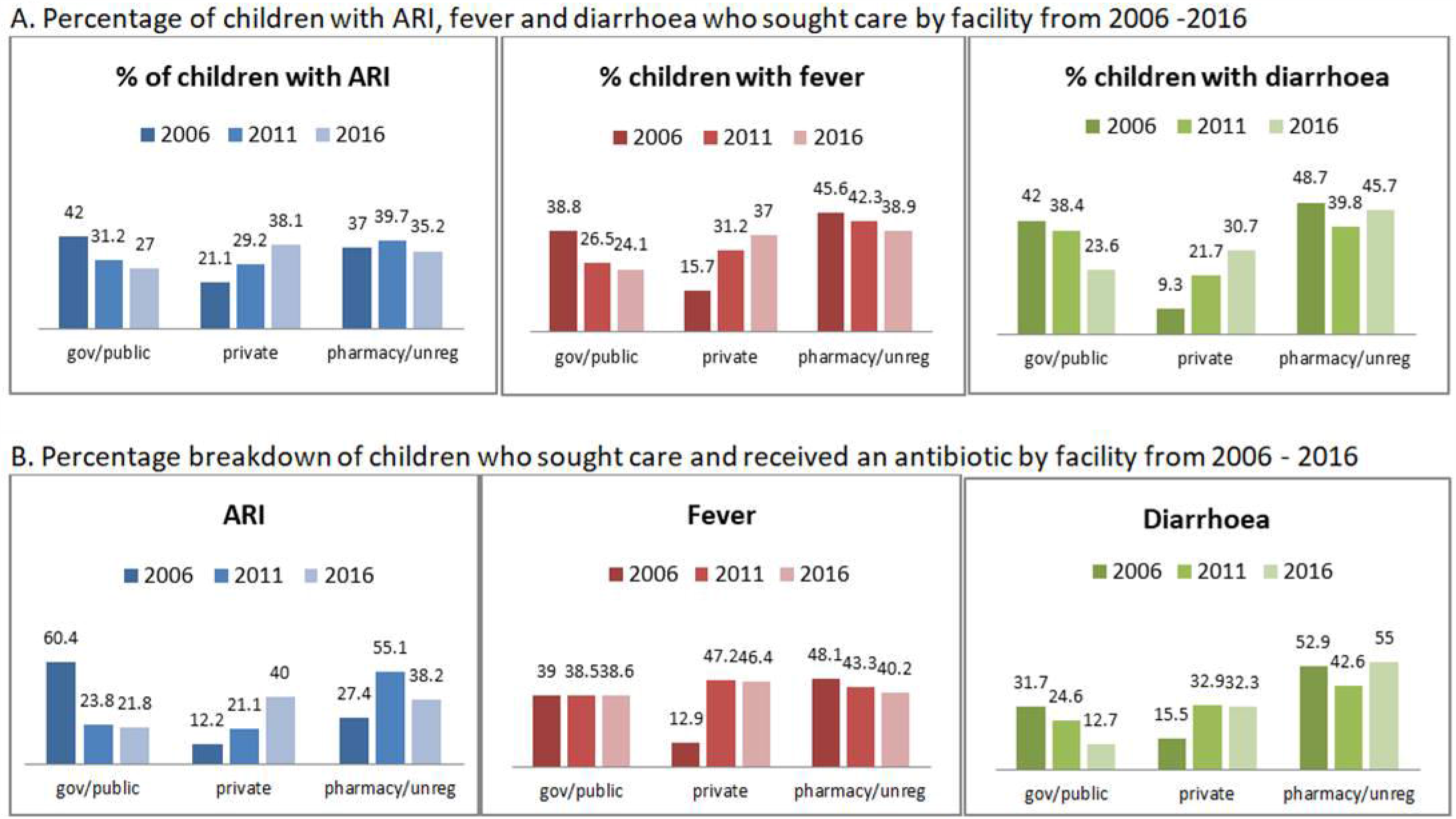

Analysis of behavioural factors demonstrated an association with clean fuel and reduced antibiotic consumption, although this was no longer significant in the multivariate analysis. Under-fives with diarrhoea, receiving the measles vaccine and all basic vaccinations in 2016, had lower rates of antibiotic consumption. Improved toilet sanitation was associated with reduced antibiotic use, whilst wasting lead to greater antibiotic use. Difficulty in accessing healthcare was associated with less antibiotic use for ARI in 2006, whereas there were higher levels of antibiotic use for fever and diarrhoea in 2016 (*Supplementary Table 4, 6 and Table 3*).

### Health-seeking behaviours

There was a decrease in the proportion of people seeking care from the public sector from 2006 – 2016 for all conditions, whilst the proportion visiting the private sector has steadily increased. The pharmacy remained a predominant healthcare provider. Unregulated facilities, including traditional healers and shopkeepers, make up a minority of the healthcare facilities initially visited (*Figure 3A*). Accordingly, the source of antibiotics reflected these changes in healthcare seeking behaviour, with the majority of those receiving antibiotics visiting a private sector facility first as opposed to the public sector in previous years. Of note, the majority of antibiotics for diarrhoea appears to be consistently coming from pharmacies since 2006 (*Figure 3B*).

### Inappropriate use of antibiotics

According to the IMCI guidelines, children with cough only should not be treated with antibiotics. From 2006 to 2011, the percentage of under-fives with cough only receiving antibiotics increased from 7% (n=15) to 12% (n=39). The figure for 2016 could not be calculated as this question was not asked. Those receiving an IMCI recommended antibiotic, specifically oral co-trimoxazole (before 2014), amoxicillin (after 2014) or IV/IM procaine penicillin, decreased from 99.9% (n=68) in 2006 to 81.6% (n=71) in 2011, and then to 50% (n=24) in 2016. The IMCI guidelines stipulate that children with diarrhoea and bloody stools (dysentery) should receive antibiotic therapy. In 2006 and 2011, only 11.5% (n=12) and 23.1% (n=23) of children with dysentery received antibiotics, whilst 75% of children who received antibiotics for diarrhoea did not meet the required indication. Again, this question was omitted in the 2016 questionnaire and could not be evaluated.

## Discussion

From our analysis, the prevalence of ARI and diarrhoea in under-fives in Nepal declined from 2006 onwards, particularly among the highest wealth quintiles. The gap between rich and poor has widened over time, which has been the global trend over the last few decades (12). It is often the case that the wealthiest groups are the first to access new services and treatments. Conversely, overall antibiotic use has been rising, reflecting increasing consumption in the low- and middle-income populations, particularly those based in rural locations. Improved access to healthcare due to economic development and the proliferation of private providers may be driving this. Falling rates of antibiotic use in the wealthier quintiles are likely to reflect concomitant reduction in disease prevalence but may also suggest improvements in maternal education (9-11), leading to increased awareness of antibiotic resistance. Even within a low-income setting, patterns of antibiotic consumption reflect global trends as described by Klein et al (1). This is not necessarily undesirable given poorer populations are often in most need of healthcare but it highlights the need for education and regulation amongst these groups to prevent inappropriate use.

Overall, clean fuel, improved sanitation, measles vaccination and nutrition are associated with reduced disease prevalence and lower antibiotic consumption. These findings emphasise the importance of optimising public health. Clean fuel use reduces indoor air pollution, which is a key contributor of acute and chronic respiratory illness (13). Improved toilet sanitation reduced diarrhoeal illness and antibiotic use but this was not seen with improved water sources, suggesting that the former could be more important in reducing enteric infections by preventing contamination of water sources in the first place. The association between measles vaccination and reduced prevalence in diarrhoea has also been demonstrated in India, Pakistan, Nigeria and the Democratic Republic of Congo (14). Measles vaccination has also been associated with increased child survival unrelated to measles prevention and reduction in hospital admissions for other infections (15, 16). Nutrition was consistently associated with reduced illness and antibiotic use in our analysis. The 2017 Global burden of disease study identified wasting as the strongest risk factor for mortality associated with pneumonia (17). Public health policy should therefore make childhood nutrition a priority.

Individuals are increasingly seeking healthcare and thus receiving antibiotics from private facilities, which is reflected by the greater number of private hospitals (364) compared to government hospitals (103) available (8). Analysis of DHS and Service Provision Assessment surveys on eight LMICs by Fink et al similarly found a higher number of under-five attendances and mean antibiotics prescribed at private health facilities versus public ones (18). Care-seeking at pharmacies has always remained high, particularly for diarrhoea. Pharmacies often provide quicker perceived solutions to more minor ailments compared to the opportunity cost of spending time being assessed in the hospital. They are likely to be a significant contributor to inappropriate antibiotic use. A study by Wachter et al. demonstrated that antibiotics were supplied inappropriately by 97% of surveyed pharmacists in Kathmandu for childhood diarrhoea (19). Simulated client studies carried out in pharmacies in India and Thailand reported inappropriate antibiotic use for diarrhoea of 40% and 52.2% respectively (20, 21). Reduced adherence to IMCI recommended antibiotics for ARI and dysentery has also been observed in other LMICs. A prospective study by Ragowski et al examining antibiotic use in children up to 2 years old in eight LMICs in South Asia, Africa and South America noted substantial variation from IMCI recommended antibiotics for ARI and in particular bloody diarrhoea (22). This could be secondary to reduced drug availability, poor knowledge of local resistance rates, financial incentives for certain antibiotics and lack of legal consequences for incorrect antibiotic prescribing (23, 24). The government needs to try and tackle unregulated antibiotic use in the private sectors but this will be challenging given prescribing laws are often ignored and difficult to implement (19). Strategies could be targeted at educating and raising awareness about antimicrobial resistance amongst these healthcare professionals.

## Limitations

As with all cross-sectional studies, the DHS data is at risk of recall bias, although non-response and selection bias was minimised by weighted data. Our analysis provides insight into the associations of demographic and behavioural variables with disease prevalence and antibiotic use but a causal relationship between exposure and outcome cannot be established, since the time sequence between exposure and outcome is not clear. Small sample subset sizes made it difficult to examine certain associations, in particular the impact of maternal smoking and breastfeeding on illness and antibiotic use. 2016 data was collected later in the year compared to 2006 and 2011 perhaps reflecting the disruption from the April 2015 earthquake. This seasonal variation along with the effect of the earthquake may have influenced trends in disease prevalence and make the 2016 data less comparable. Lastly, the data only provided a limited perspective on the ‘appropriate’ use of antibiotics, as neither clinical information nor microbiology results were available.

## Conclusion

The results of our study highlight the importance of the following specific primary strategies that were proposed in the Situation Analysis conducted with the Global Antibiotic Resistance Partnership to improve antibiotic use: reduce the need for antibiotics by improving public health (in particular sanitation, nutrition and reduced indoor air pollution), rationalize antibiotic use in the community and educate health professionals, policy makers and the public on sustainable antibiotic use (7). These measures need to be balanced with ensuring those who need antibiotics the most can still access them.

## Data Availability

Data sources are available from the Demographic Health Survey (DHS) website.

https://dhsprogram.com/What-We-Do/survey-search.cfm?pgtype=main&SrvyTp=country

